# Uptake and retention in HIV care among pregnant and postpartum women living with HIV under different eras of vertical transmission prevention policies in sub-Saharan Africa: a systematic review and meta-analysis

**DOI:** 10.64898/2026.04.02.26350030

**Authors:** Nelly Jinga, Candice Hwang, Laura Rossouw, Kate Clouse, Cornelius Nattey, Bernard Mbwele, Nkosinathi Ngcobo, Molly Beestrum, Mark D. Huffman, Matthew P. Fox, Mhairi Maskew

## Abstract

**Objectives:** This systematic review and meta-analysis (2010–2025) examines changes in uptake and retention rates among pregnant and postpartum women with HIV in sub-Saharan Africa as countries adopted Option B+ for preventing vertical transmission.

**Design and data sources:** We searched PubMed, Embase, Cochrane Library, Scopus, and African Index Medicus from 10/2021–05/2025 for eligible studies that measured HIV care uptake or retention for pregnant/postpartum women under prevention policies before or during Option B+. Study designs were limited to cohort, case-control, cross-sectional, or interventional studies. Exclusions were white papers, commentaries, modeling, cost-effectiveness, and qualitative studies.

**Data extraction and synthesis:** Outcomes were (i) HIV care uptake defined as initiation of ART during pregnancy or prior to initial antenatal care (ANC) visit and (ii) proportion of women retained in HIV care as defined by study authors after ART initiation (or entry to antenatal care). These were synthesized in meta-analyses stratified by policy era (pre-Option B+ vs. Option B+) at different times for different countries. Comparisons between policy eras were made using relative risk with a 95% confidence interval. Pooled retention estimates at 6- and 12-months post ART initiation used crude relative risks (RR) with 95% confidence intervals (CI).

**Results:** Among 4,752 articles, 82 from 17 countries were included; 60 reported HIV care uptake, 31 reported retention outcomes. Pooled HIV uptake rose by 8% (RR=1.08; 95% CI:1.06-1.09) and pooled retention in HIV care rose by 46% (RR=1.46; 95% CI:1.41-1.51) after Option B+ implementation. Pooled estimates of retention in care were 36.9% (95% CI: 13.9%, 59.9%) at 6 months post ART initiation before the implementation of Option B+ and 72.7% (95% CI: 66.3%, 79.1%) after implementation.

**Conclusion:** HIV care uptake and retention improved after Option B+ implementation in 15 countries reporting results, but retention remains suboptimal for meeting UNAIDS 95-95-95 targets.

**Strengths and Limitations:**

- Large number of included studies from diverse population groups.
- This review includes only published studies, which may limit generalizability.
- Reviews included 15 countries of sub-Saharan Africa; therefore, results may not be generalizable to other parts of sub-Saharan Africa or other regions.
- Our review may overestimate effects after funnel plot and Egger’s test, which may cause publication bias towards studies with significant or favourable findings and the possibility of exclusion of publications with null findings in the published literature.
- The review consists of studies that may have only included those who got onto ART resulting in overestimating the uptake of HIV care proportions.

## INTRODUCTION

Major gains have been made in the prevention of vertical transmission (PVT) of HIV over the past two decades. Globally, the percentage of pregnant women living with HIV who received antiretroviral therapy (ART) for PVT rose from 45% in 2010 to 84% in 2023.^1^ This has been driven by the progression of World Health Organization (WHO) recommendations for prevention of mother-to-child transmission (PMTCT) of HIV protocols. In 2010, WHO guidelines outlined Option A and B (see Table 1 for definitions) where ART for prophylaxis was offered temporarily during pregnancy and breastfeeding.^2^ The 2013 WHO guidelines expanded ART access by introducing Option B+, where pregnant women were offered lifetime ART regardless of CD4 count. Following the release of the 2013 guidelines, many countries moved to implement Option B+, and by 2015, the WHO recommended only Option B+ moving forward.^3^

**Table 1.** Definitions of Option A, B, and B+ policies.^2^.

|  | Woman receives: |  | Infant receives: |
| --- | --- | --- | --- |
|  | Treatment<br>(for CD4 count<br>≤350 cells/mm<sup>3</sup>) | Prophylaxis<br>(for CD4 count<br>>350 cells/mm<sup>3</sup>) |  |
| Option A | Three ARVs starting as soon as diagnosed, continued for life | *Antepartum:* AZT starting as early as 14 weeks gestation<br>*Intrapartum:* at onset of labor, sdNVP and first dose of AZT/3TC<br>*Postpartum:* daily AZT/3TC through 7 days postpartum | Daily NVP from birth through 1 week beyond complete cessation of breastfeeding; or, if not breastfeeding or if mother is on treatment, through age 4–6 weeks |
| Option B | Three ARVs starting as soon as diagnosed, continued for life | Triple ARVs starting as early as 14 weeks gestation and continued intrapartum and through childbirth if not breastfeeding or until 1 week after cessation of all breastfeeding | Daily NVP or AZT from birth through age 4–6 weeks regardless of infant feeding method |
| Option B+ | Same for treatment and prophylaxis: |  | Daily NVP or AZT from birth through age 4–6 weeks regardless of infant feeding method |
|  | Regardless of CD4 count, three ARVs starting as soon as diagnosed, continued for life |  |  |

Abbreviations: ARV = antiretroviral drugs, AZT = Zidovudine, 3TC = Lamivudine, NVP = Nevirapine

Option B+ has been one of the key developments in countries’ PVT programmes worldwide. This set of guidelines gave hope for achieving universal access to HIV care and treatment and ending AIDS as a public health threat.^4^ However, there have been challenges in implementing Option B+ leading to low retention in care and poor ART adherence during the prenatal and postpartum periods.^5, 6, 7^

In 2020, UNAIDS released a series of interim targets for 2025 that need to be achieved to reach the 2030 HIV Sustainable Development Goal targets. The UNAIDS 2025 targets include: 95% of women living with HIV on ART before their current pregnancy, 95% of pregnant women living with HIV achieving viral suppression before delivery, and 95% of breastfeeding women living with HIV achieving viral suppression at 6-12 months postpartum.^8^ Despite substantial progress in expanding ART access and reducing vertical transmission of HIV, persistent challenges in uptake and retention in HIV care among pregnant and postpartum women threaten progress toward elimination goals. Evidence to date remains fragmented, with studies differing in design, setting, and measurement of outcomes. Moreover, while numerous evaluations have focused on ART initiation during pregnancy, fewer have systematically synthesized data on retention and viral suppression in the postpartum period—key determinants of sustained maternal and infant health outcomes. A comprehensive synthesis of recent evidence is therefore needed to assess how uptake and retention have evolved under Option B+ in South Africa and the broader sub-Saharan African context, to identify persistent barriers, and to benchmark current progress against the UNAIDS 2025 targets. This systematic review aims to summarize updated data on uptake and retention in HIV care for pregnant and postpartum women living with HIV in sub-Saharan Africa from 2010 to the present, assess changes in uptake and retention rates as countries transitioned to Option B+, and compare the published uptake and retention rates to the UNAIDS 2025 targets.

## METHODS

We used the Preferred Reporting Items for Systematic Reviews and Meta-Analyses (PRISMA) 2020 guidelines.^9^ We conducted a systematic review of peer-reviewed publications and published conference abstracts that reported on uptake or retention in HIV care among pregnant and postpartum women living with HIV in sub-Saharan Africa. We included studies of women who received care under any of the following PVT policies: Option A, Option B and Option B+ (Table 1). The review was prospectively registered on the International Prospective Register of Systematic Reviews (PROSPERO CRD4202129012) (Supplementary File 1).

### Search strategy, study selection and data extraction

We searched the following databases: PubMed; Embase, Cochrane Library, Scopus, and African Index Medicus. Abstracts from the International AIDS Conference, International AIDS Society (IAS) Conference on HIV Science, and Conference on Retroviruses and Opportunistic Infections (CROI) are indexed in Scopus and were captured in the search.

We developed a search string with the assistance of a medical librarian (MB). Search syntax included terms related to: 1. HIV and pregnancy or the postpartum period; 2. Antiretroviral therapy; 3. Uptake or retention outcomes; and 4. Africa or any sub-Saharan African country. Searches were limited to English language publications. The full search strategy is detailed in Appendix Table 1. We limited our search to articles published after Jan 1, 2010, to align with the WHO recommendations published in 2010 that introduced Option A and B and to allow time for guideline adoption.

We included studies that measured uptake or retention in HIV care outcomes for pregnant and post-partum women in the context of a PVT policy implemented in the country. Study types included cohort, case-control, cross-sectional, or interventional studies. In interventional studies where retention or uptake were the target of intervention, only data from the control arm were included. White papers, commentaries, modeling studies, cost-effectiveness studies, and case studies were excluded. Studies could involve any type of facility or provider. Qualitative studies were excluded unless there were specific data on uptake or retention outcomes. Existing systematic reviews were evaluated for additional, non-duplicate references. While published protocols with no data were excluded, for the studies that met the inclusion criteria we searched for study results to evaluate for inclusion. As “PMTCT” was the term widely used for many years, it was part of our search strategy; however, in this paper, we use the person-first language recommendation of “PVT” since “PMTCT” has potentially negative connotations related to implied blame and may minimize male involvement.^10^

All identified references were imported into EndNote, where deduplication occurred. The Rayyan web application was used to manage the studies and track inclusion or exclusion of studies. Two independent reviewers were involved in each phase of review, including title and abstract screening, full-text screening, and data extraction. Title and abstract screening included a pilot test to assess and ensure that reviewers achieved over 90% agreement on inclusion based on the eligibility criteria. CH and NJ led the review process, and the second reviewers included BM, CN NN, LR and MM, with studies randomly distributed among them. Disagreements were resolved through discussion to achieve consensus or input from MM to achieve consensus. Data extraction used a predetermined template that captures all relevant variables.

We extracted the following data items from all studies, country, district or locality, facility type or setting, study year(s), length of patient follow-up, PVT protocol(s) that were used, study sample size, patient characteristics (mean age, gravida, clinical indicators of disease stage such as WHO stage, other co-morbidities), study design (e.g., cohort, case control, cross-sectional, interventional), descriptions of routine care, including whether the study was designed to influence uptake or retention.

### Outcomes and analysis

Our outcomes were twofold: 1) uptake of HIV care, defined as initiation of ART during pregnancy or prior to initial antenatal care (ANC) visit, and 2) proportion of women retained in HIV care as defined by study authors after ART initiation (or entry to antenatal care). For uptake of antenatal HIV care, the ART regimen at initiation had to, at minimum, meet Option A criteria for treatment and prophylaxis. For retention, patients reported as non-adherent to ART but attending clinic appointments (or meeting other study-defined criteria for retention) were counted as retained as were patients who were known to have transferred care to another facility. Retention was reported at the 6-, 12- and 24-months’ time point after ART start.

Secondary outcomes considered included median gestational age at initiation of ART during pregnancy, median gestational age at initial ANC visit, mean CD4 count at initial ANC visit, mean viral load at initial ANC visit and mean viral load (or categorical suppressed vs not suppressed) at delivery.

We commenced our analysis by describing each study’s characteristics, including population, context, and outcomes. The rates of HIV care uptake and retention in HIV care prior to Option B+ and during Option B+ era were compared. The stratification by PVT era was done according to which PVT policy was in place at the time the study was conducted in that country, and not according to year. Retention at 6- and 12-months was also summarized using meta-analysis without policy era stratification. Pooled estimates of the proportion of women retained in care at 6- and 12-months were generated using crude relative risks (RR) with 95% confidence intervals (CI) also stratified by policy era. Retention estimates and their 95% confidence intervals were plotted using forest plots. Sensitivity was assessed with the I-squared statistic. The pooled estimates accounted for this heterogeneity using a random effects regression with a Freeman and Tukey arcsine transformation.

### Critical appraisal and risk of bias assessment

A thorough evaluation of quality, encompassing critical appraisal and risk of bias, was independently conducted using the Joanna Briggs Institute (JBI) critical appraisal checklist tailored for studies reporting prevalence data. The critical appraisal form was used to systematically assess several aspects of quality, including sample representativeness of the target population, the recruitment method for study participants, adequacy of sample size, description of study subjects and setting, method of data analysis, adherence to standard criteria for measurement, reliability of outcome measurement, appropriateness of statistical analysis, identification and handling of significant confounding factors, and identification of subpopulations using objective criteria. A pilot of risk of bias assessment was done by CH and NN with 20% of the included studies. The remaining risk of bias assessment was completed by NN & NJ. An Egger’s test and a funnel plot were applied to assess publication bias.

### Patient and Public Involvement

Patients and the public were not involved in this study.

## RESULTS

### Sources identified

The primary search was conducted from October 2021 to May 2025, and 4752 articles were reviewed with 328 reports included in full text screening. Eighty-two full text reports on estimation of retention in HIV care among pregnant and postpartum women living with HIV with implementation of different PVT policies were included from 17 sub-Saharan countries, 59 articles reported results related to HIV care uptake, including 26 that reported both and 31 articles reported results related to retention. Twenty-five articles included results before Option B+ implementation, 57 articles included results after Option B+ implementation, and 2 articles included results during both eras (Figure 1).

**Figure 1.**
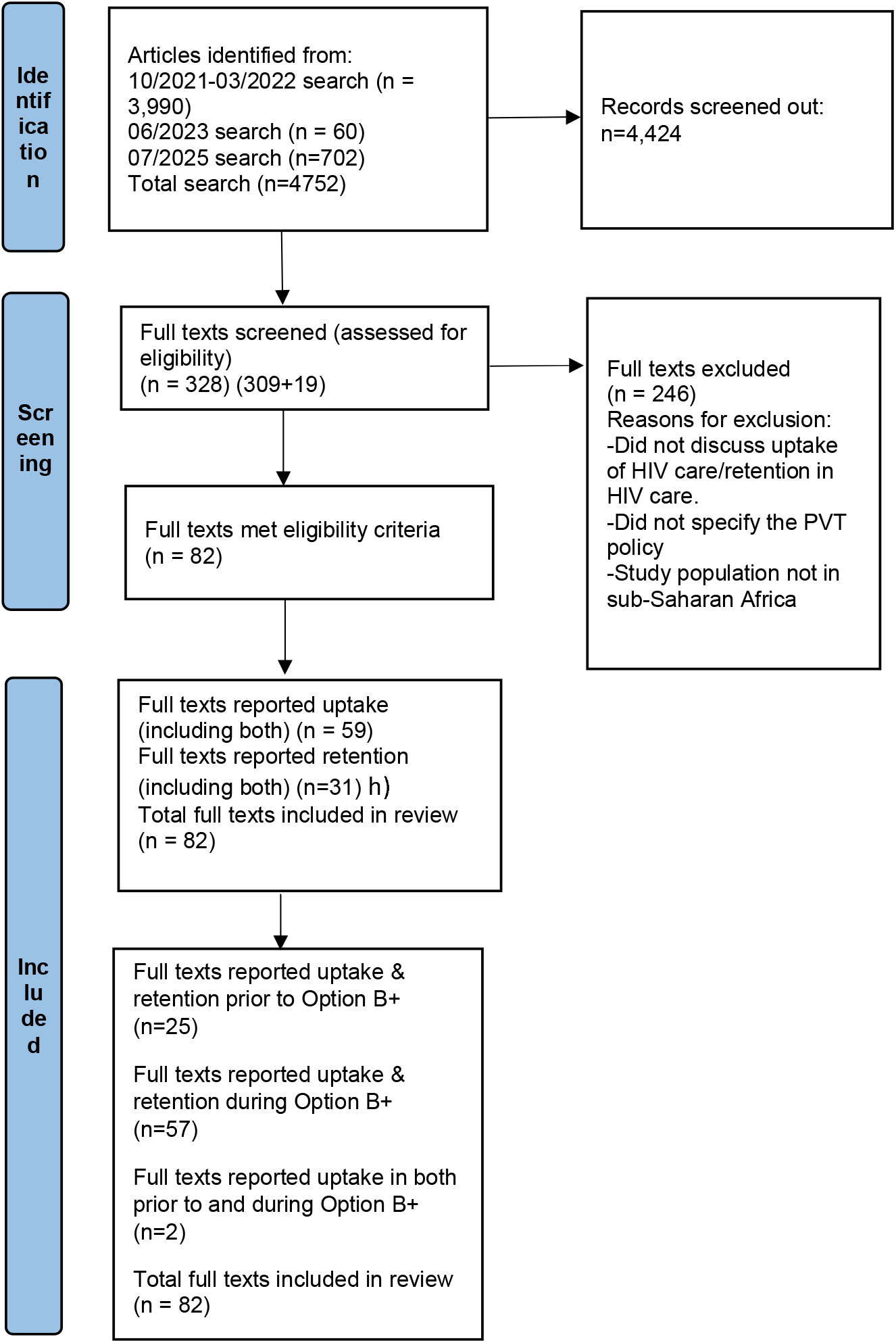
PRISMA flowchart of the search protocol for studies reporting on uptake and retention of prevention of vertical transmission of HIV from 2010-2023 in sub-Saharan Africa resulting in 61 studies included in the systematic review.

Table 2 summarizes the characteristics of all the 61 studies included in the analysis. Sample sizes in the studies examined varied from 49 to 32,015 participants. Among the studies that included CD4 counts, only two studies reported women initiating antenatal care with a CD4 count below 200 cells/mm^3^. Among studies that reported mean age at initiation of antenatal care, 85% (46/54) reported a mean age that was below 30 years and among studies that reported gestational age at antenatal care initiation 31% (8/26) reported that the average gestational age was 20 weeks. The mean study follow-up period was 2 years and the maximum follow-up period was 8 years.

**Table 2.** Characteristics of studies included in the review (n=61) of studies reporting on uptake and retention of prevention of vertical transmission (PVT) of HIV in sub-Saharan Africa from 2010-2023.

| Author and year | PVT policy | Study Design | Reported outcomes | Start year of study period | End year of study period | Sample size | Age (mean) | Gestation al age at initial ANC visit | CD4 count at initial ANC visit (median) |
| --- | --- | --- | --- | --- | --- | --- | --- | --- | --- |
| Abrams et al. (2019)<sup>11</sup> | Option A | Non-randomized interventional | Retention in care, proportion of women initiating ART during pregnancy, time from first ANC to ART initiation | 2013 | 2015 | 1296 | 26.7 | 20 | 412 |
| Clouse et al. 2013<sup>12</sup> | Option A | Cohort | Retention in care during ANC and postpartum. | 2010 | 2010 | 273 | 27 | 26 | 357 |
| Dinh et al. 2018<sup>13</sup> | Option A | Cohort | ART uptake, risk of mother to child transmission. | 2013 | 2014 | 1172 |  |  |  |
| Fatti et al. 2016<sup>14</sup> | Option A | Cohort | Time from the first antenatal visit to initiating ART, risk of not initiating antenatal, proportion of infants stillborn and vertical HIV transmission. | 2009 | 2012 | 841 | 27.4 |  | 361 |
| Giuliano et al. 2016<sup>15</sup> | Option A | Cohort | Loss to follow-up. | 2008 | 2009 | 311 | 27 | 23 | 339 |
| Granato et al. 2016<sup>16</sup> | Option A |  | Infant ART uptake, retention in care. | 2011 | 2012 | 1741 |  |  |  |
| Hussain et al. 2011<sup>17</sup> | Option A | Cross sectional | Uptake of HIV care. | 2010 | 2010 | 630 |  |  | 285 |
| Kamuyango et al. 2014<sup>18</sup> | Option A | Cohort | Treatment outcomes one year after ART initiation | 2008 | 2010 | 102 | 29 |  | 231 |
| Kim et al. 2015<sup>19</sup> | Option A | Pre/post quasi-experimental study | Uptake of PMTCT, antenatal outcomes among women enrolled into PMTCT. | 2009 | 2011 | 1129 | 26.9 |  |  |
| Vogt et al. 2015<sup>20</sup> | Option A | Cohort | Retention in care. | 2010 | 2013 | 1278 | 27 |  | 394 |
| Van Schalkwyk et al. 2018<sup>21</sup> | Pre-Option A | Cohort | ART uptake. | 2008 | 2010 | 250 | 28 | 20 | 176 |
| Ross et al. 2018<sup>22</sup> | Option A | Cohort | Retention in care, and loss to care. | 2007 | 2013 | 1497 | 30 |  | 349 |
| Phillips et al. 2014<sup>23</sup> | Option A | Secondary analysis of prospective cohort study. | Disengagement from care, non-adherence. | 2011 | 2012 | 358 | 28 |  | 233 |
| Aliyu et al. 2015<sup>24</sup> | Option A/B | Cohort | Difference in ART initiation and retention by sex among enrolled clients and ART initiators between June 2009 and September 2013. | 2009 | 2013 | 273 | 27 |  | 306 |
| Aliyu et al. 2016<sup>25</sup> | Option B | Randomized controlled trial | Proportion of eligible women who initiated antiretroviral medications for PMTCT and the proportion of HIV-infected women and their infants retained in care. | 2013 | 2014 | 197 | 28 | 20 | 314 |
| Escamilla et al. 2015<sup>26</sup> | Option B | Cross sectional | Uptake of PMTCT, uptake of combination antiretroviral regimens. | 2011 | 2011 | 254 |  |  |  |
| Hwang et al. 2024<sup>27</sup> | Option B | Cohort | Uptake and engagement in antenatal and HIV care | 2013 | 2014 | 5483 | 27 | 21 |  |
| Sam-Agudu et al. 2017<sup>28</sup> | Option B | Non-randomized interventional | Retention in care, maternal viral suppression. | 2014 | 2015 | 497 |  |  |  |
| Mnyani et al. 2020<sup>29</sup> | Option B | Retrospective study of routinely collected PMTCT data | Proportion first antenatal visit, uptake of HIV testing, uptake of ART initiation, uptake of CD4 testing, infant uptake of HIV test and proportion testing HIV positive. | 2013 | 2014 | 12146 |  |  |  |
| Ongaki et al. 2019<sup>30</sup> | Option B | Retrospective record review | Uptake of PMTCT services | 2015 | 2016 | 230 | 26 |  |  |
| Wanyenze et al. 2018<sup>31</sup> | Option B | Cohort | Uptake of PMTCT services | 2013 | 2013 | 127 | 31.7 | 15.5 |  |
| Myburgh et al. 2017<sup>32</sup> | Option B | Cohort | Adherence to the PMTCT policy. | 2013 | 2014 | 100 | 32 |  | 412 |
| Myer et al. 2012<sup>33</sup> | Before B+ | Randomized controlled trial | Retention in care and virologic suppression, loss to follow-up | 2003 | 2010 | 490 | 27 | 28 | 142 |
| Myer et al. 2018<sup>34</sup> | Straddles Option A to B+ transition | Cohort | Retention in ART care and viral suppression 12-months postpartum. | 2013 | 2014 | 471 | 28.6 | 21 | 354 |
| Abrams et al. (2019)<sup>11</sup> | Option B+ | Non-randomized interventional | Retention in care, proportion of women initiating ART during pregnancy, time from first ANC to ART initiation | 2013 | 2015 | 1051 | 26.2 | 20 | 391 |
| Ahoua et al. (2020)<sup>35</sup> | Option B+ | Cohort | Attrition and Non-retention | 2013 | 2017 | 31186 | 25 |  | 386 |
| Akama et al. (2019)<sup>36</sup> | Option B+ | Cohort | Uptake of viral load testing, retention in care, | 2014 | 2016 | 165 |  |  |  |
| Atanga et al. 2017<sup>37</sup> | Option B+ | Cohort | HCT uptake, ART uptake, Retention in care. | 2013 | 2014 | 404 | 27.9 |  | 420 |
| Chaka et al. 2019<sup>38</sup> | Option B+ | Cohort | PMTCT Option B+ uptake. | 2013 | 2016 | 248 | 26.8 | 18 | 528 |
| Chan et al. 2016<sup>39</sup> | Option B+ | Cohort | HIV testing and Uptake of ART, retention on ART. | 2011 | 2012 | 914 |  |  |  |
| Chimwaza et al. 2021<sup>40</sup> | Option B+ | Cohort | ART retention. | 2018 | 2018 | 388 | 29 |  |  |
| Dzangare et al. 2016<sup>41</sup> | Option B+ | Cohort | Time from HIV diagnosis to initiation of Option B+, Treatment outcomes, attrition from care. | 2014 | 2014 | 282 |  |  |  |
| Erlwanger et al. 2017<sup>42</sup> | Option B+ | Randomized controlled trial | Retention in care. | 2014 | 2015 | 1113 | 26 |  |  |
| Etoori et al. 2018<sup>43</sup> | Option B+ | Cohort | Time from first ANC visit to ART initiation, time to loss to follow-up or death, retention in care, HIV mother to infant transmission, early infant diagnosis. | 2013 | 2014 | 665 | 26 | 22 | 387 |
| Ford et al. 2017<sup>44</sup> | Option B+ | Implementation study | Retention on ART. | 2013 | 2015 | 386 |  |  |  |
| Gamell et al. 2017<sup>45</sup> | Option B+ | Cohort | Uptake of PMTCT, MTCT rate | 2014 | 2015 | 124 |  |  |  |
| Girma et al. 2017<sup>46</sup> | Option B+ | Cohort | Retention in PMTCT care, maternal adherence to ART, infant ART uptake. | 2012 | 2015 | 494 | 28 |  | 387 |
| Haas et al. 2016<sup>47</sup> | Option B+ | Cohort | Retention on ART. | 2011 | 2012 | 29768 | 30 |  |  |
| Hosseini pour et al. 2017<sup>48</sup> | Option B+ | Randomized controlled trial | Retention in care. | 2013 | 2014 | 447 |  |  |  |
| Joseph et al. 2017<sup>49</sup> | Option B+ | Randomized controlled trial | Retention on ART. | 2014 | 2015 | 547 | 27 | 21 |  |
| Kamuyango et al. 2014<sup>18</sup> | Option B+ | Cohort | Treatment outcomes one year after ART initiation. | 2011 | 2012 | 190 | 27 |  | 558 |
| Kim et al. 2015<sup>19</sup> | Option B+ | Pre/post quasi-experimental study | Uptake of PMTCT, antenatal outcomes among women enrolled into PMTCT. | 2011 | 2013 | 1417 | 27.8 |  |  |
| Kiwanuka et al. 2018<sup>50</sup> | Option B+ | Cohort | Retention in care and LTFU. | 2013 | 2015 | 518 |  |  |  |
| van Lettow et al. 2018<sup>51</sup> | Option B+ | Cross sectional | Uptake of maternal ART. | 2014 | 2016 | 3233 |  |  |  |
| Llenas-García et al. 2016<sup>52</sup> | Option B+ | Cohort | Retention in care. | 2013 | 2014 | 243 | 24 |  | 495 |
| Lyatuu et al. 2021<sup>53</sup> | Option B+ | Cohort | Viral suppression. | 2014 | 2016 | 15586 |  |  |  |
| Mandewo et al. 2020<sup>54</sup> | Option B+ | Cross sectional | HIV testing uptake and ART initiation for pregnant women. | 2014 | 2018 | 6097 |  |  |  |
| Tweya et al. 2014<sup>55</sup> | Option B+ | Cohort | Loss to follow-up. | 2011 | 2013 | 2930 | 26 |  |  |
| Mazuguni et al. 2019<sup>56</sup> | Option B+ | Cohort | Loss to follow-up. | 2014 | 2015 | 468 | 29.29 |  |  |
| Mitiku et al. 2016<sup>57</sup> | Option B+ | Cohort | Loss to follow-up. | 2013 | 2015 | 346 | 26 | 20 | 460 |
| Tenthani et al. 2014<sup>58</sup> | Option B+ | Cohort | Loss to follow-up. | 2011 | 2012 | 11534 |  |  |  |
| Schwartz et al. 2015<sup>59</sup> | Option B+ | Cohort | Time from HIV diagnosis to ART initiation, death, transfer out and stopped ART. | 2013 | 2013 | 50 | 29 | 21 | 434 |
| Reidy et al. 2019<sup>60</sup> | Option B+ | Cohort | Maternal and/or infant engagement in care, use of ART by women. | 2013 | 2014 | 434 | 24 |  |  |
| Pricilla et al. 2018<sup>61</sup> | Option B+ | Cohort | ART initiation during pregnancy, infant HIV transmission rate. | 2013 | 2016 | 2371 | 29 |  |  |
| Price et al. 2014<sup>62</sup> | Option B+ | Retrospective cohort study | Uptake of PMTCT. | 2011 | 2013 | 63 | 25 |  |  |
| Phiri et al. 2017<sup>63</sup> | Option B+ | Cohort | ART uptake, retention in care. | 2013 | 2014 | 1269 | 27 |  |  |
| Phillips et al. 2018<sup>64</sup> | Option B/B+ | Randomized controlled trial | Linkage to care. | 2013 | 2014 | 617 | 29 | 21 | 345 |
| Myer et al. 2015<sup>65</sup> | Option B+ | Randomized controlled trial | Proportion of women completing different steps within the PMTCT 'cascade'. | 2010 | 2013 | 19432 |  |  | 390 |
| Muhumuza et al. 2017<sup>66</sup> | Option B+ | Cohort | Retention in care. | 2013 | 2013 | 2169 | 25 |  | 524 |
| Abuogi et al. 2022<sup>67</sup> | Option B+ | Cluster randomised control | Retention in care and ART adherence, viral suppression and infant outcomes. | 2015 | 2017 | 1331 | 28 | 24 |  |
| Geremew et al. 2023<sup>68</sup> | Option B+ | Retrospective follow-up study | Loss to follow-up from option B+ PMTCT service | 2013 | 2021 | 365 |  |  |  |
| Humphrey et al. 2022<sup>69</sup> | Option B+ | Retrospective cohort study | Retention in care, Loss to follow-up. | 2015 | 2019 | 856 |  | 20 |  |
| Teasdale et al. 2022<sup>70</sup> | Option B+ | Evaluation study | Incidence rate of LTFU in the first six months after enrollment in ANC services. | 2017 | 2019 | 543 |  | 21 |  |
| Chagomerana et al. 2023<sup>71</sup> | Option B+ | Prospective cohort study | Retention in care and clinical and laboratory-confirmed outcomes. | 2015 | 2019 | 299 | 27 | 22.1 |  |
| Humphrey et al. 2024<sup>72</sup> | Option B+ | Prospective cohort study | Retention in HIV care | 2018 | 2018 | 333 | 31 | 23 |  |
| Naidoo et al. 2023<sup>73</sup> | Option B+ | Retrospective record review | Mother to Child Transmission (MTCT) rate | 2019 | 2020 | 658 |  |  |  |
| Phelanyane et al. 2023<sup>74</sup> | Option B+ | Cohort | Vertical transmission during pregnancy & postpartum | 2017 | 2017 | 2549 |  |  |  |
| Ameyaw et al. 2024<sup>75</sup> | Option B+ | Cross sectional | Uptake of ART, Retention | 2023 | 2023 | 176 | 32 |  |  |
| Giovenco et al. 2025<sup>76</sup> | Option B+ | Prospective cohort study | Retention in HIV care | 2020 | 2022 | 375 |  | 20 |  |
| Azanaw et al. 2023<sup>77</sup> | Option B+ | Retrospective follow-up | Retention in HIV care | 2013 | 2019 | 403 | 27.6 |  | 419.1 |
| Slogrove et al. 2024<sup>78</sup> | Option B+ | Retrospective cohort study | Uptake of HIV care. | 2018 | 2019 | 32015 | 26.9 |  | 425 |
| Camara et al. 2024<sup>79</sup> | Option B+ | Retrospective cohort study | Uptake of HIV care. | 2019 | 2022 | 385 |  |  |  |
| Amone et al. 2024<sup>80</sup> | Option B+ | Randomized controlled trial | Retention in HIV care | 2016 | 2020 | 270 |  |  |  |
| Bengtson et al. 2024<sup>76</sup> | Option B+ | Prospective cohort study | Disengagement from HIV care | 2020 | 2022 | 402 | 30 | 20 |  |
| Greenberg et al. 2025<sup>81</sup> | Option B+ | Cluster randomised controlled trial | Adherence to HIV therapy | 2016 | 2017 | 304 | 28 | 21 |  |
| Mutale et al. 2025<sup>82</sup> | Option B+ | Randomized controlled trial | Retention in HIV care | 2020 | 2021 | 49 | 27 | 24 |  |
| Njah et al. 2023<sup>83</sup> | Option B+ | Retrospective | Retention in HIV care | 2015 | 2016 | 560 | 28 | 21 |  |
| Odongo et al. 2023<sup>84</sup> | Option B+ | Longitudinal analysis | Retention in HIV care | 2013 | 2018 | 368 | 29.7 |  |  |
| Phillips et al. 2024<sup>85</sup> | Option B+ | Cohort | Disengagement from HIV care | 2013 | 2019 | 6680 | 28 |  | 325 |
| Young et al. 2024<sup>86</sup> | Option B+ | Retrospective cohort study | LTFU, ART retention, adherence and viral suppression | 2013 | 2017 | 1535 | 33 |  |  |
| Tolossa et al. 2020<sup>87</sup> | Option B+ | Cross sectional | LTFU from PMTCT services | 2013 | 2018 | 330 |  |  |  |

**Abbreviations:** ANC: antenatal care; ART: antiretroviral treatment; HCT: HIV counselling and testing; LTFU: Loss to follow-up; PMTCT: Prevention of mother to child transmission of HIV; PVT: Prevention of vertical transmission of HIV; NR: Not reported

### Uptake of HIV care

The first outcome considered was uptake of HIV care. In total, 59 studies (72%) reported uptake of HIV care data (Table 3) among pregnant and postpartum women with HIV in sub-Saharan Africa. Most studies (78%, n= 46/59) looked at uptake of HIV care among pregnant and postpartum women during the Option B+ era. Sample sizes varied widely, from 49 to 32,015 participants. More than 60% of studies had an HIV care uptake of 100%; however, these studies likely are biased since they only included those who started ART. The median uptake of HIV care increased from 90% (IQR: 76-100) before Option B+ to 100% (IQR: 82-100) after implementation. The pooled estimate of uptake of HIV care prior to Option B+ was 76% versus 82% after implementation of Option B+ (RR=1.08, 95% CI=1.06-1.09).

**Table 3.** Uptake of HIV care among studies of prevention of vertical transmission (PVT) of HIV in sub-Saharan Africa from 2010-2023 by PVT policy era.

| Author | PVT policy | Number initiated on ART | Sample size | Uptake (%) |
| --- | --- | --- | --- | --- |
| **Pre-Option B+ era** |  |  |  |  |
| Giuliano et al. 2016<sup>15</sup> | Option A | 311 | 311 | 100 |
| Van Schalkwyk et al. 2013<sup>21</sup> | Pre-Option A | 250 | 250 | 100 |
| Kamuyango et al. 2014<sup>18</sup> | Option A | 102 | 102 | 100 |
| Ross et al. 2018<sup>22</sup> | Option A | 1497 | 1497 | 100 |
| Philips et al. 2014<sup>23</sup> | Option A | 358 | 358 | 100 |
| Sam-Agudu et al. 2017<sup>28</sup> | Option B | 497 | 497 | 100 |
| Wanyenze et al. 2018<sup>31</sup> | Option B | 103 | 127 | 81 |
| Phillips et al. 2018<sup>7</sup> | Shifted to B+ mid-study | 485 | 617 | 79 |
| Myer et al. 2012<sup>33</sup> | Option B | 382 | 490 | 78 |
| Aliyu et al. 2015<sup>24</sup> | Option A/B | 213 | 273 | 78 |
| Myer et al. 2018<sup>34</sup> | Straddles Option A to B+ transition | 356 | 471 | 76 |
| Kim et al. 2015<sup>19</sup> | Option A | 225 | 1129 | 20 |
| Clouse et al. 2013<sup>12</sup> | Option A | 93 | 273 | 34 |
| **Option B+ era** |  |  |  |  |
| Ahoua et al. (2020)<sup>35</sup> | Option B+ | 31186 | 31186 | 100 |
| Akama et al. (2019)<sup>36</sup> | Option B+ | 165 | 165 | 100 |
| Ford et al. 2017<sup>44</sup> | Option B+ | 386 | 386 | 100 |
| Gamell et al. 2017<sup>45</sup> | Option B+ | 124 | 124 | 100 |
| Chimwaza et al. 2021<sup>40</sup> | Option B+ | 388 | 388 | 100 |
| Haas et al. 2016<sup>47</sup> | Option B+ | 29768 | 29768 | 100 |
| Erlwanger et al. 2017<sup>42</sup> | Option B+ | 1113 | 1113 | 100 |
| Hosseini pour et al. 2017<sup>48</sup> | Option B+ | 447 | 447 | 100 |
| Joseph et al. 2017<sup>49</sup> | Option B+ | 547 | 547 | 100 |
| Kamuyango et al. 2014<sup>18</sup> | Option B+ | 190 | 190 | 100 |
| Kiwanuka et al. 2018<sup>50</sup> | Option B+ | 518 | 518 | 100 |
| Llenas-García et al. 2016<sup>52</sup> | Option B+ | 243 | 243 | 100 |
| Lyatuu et al. 2021<sup>53</sup> | Option B+ | 15586 | 15586 | 100 |
| Tweya et al. 2014<sup>55</sup> | Option B+ | 2930 | 2930 | 100 |
| Mazuguni et al. 2019<sup>56</sup> | Option B+ | 468 | 468 | 100 |
| Mitiku et al. 2016<sup>57</sup> | Option B+ | 346 | 346 | 100 |
| Muhumuza et al. 2017<sup>66</sup> | Option B+ | 2169 | 2169 | 100 |
| Abuogi et al. 2022<sup>67</sup> | Option B+ | 1331 | 1331 | 100 |
| Geremew et al. 2023<sup>68</sup> | Option B+ | 365 | 365 | 100 |
| Humphrey et al. 2022<sup>69</sup> | Option B+ | 856 | 856 | 100 |
| Teasdale et al. 2022<sup>70</sup> | Option B+ | 543 | 543 | 100 |
| Chagomerana et al. 2023<sup>71</sup> | Option B+ | 299 | 299 | 100 |
| Humphrey et al. 2024<sup>88</sup> | Option B+ | 333 | 333 | 100 |
| Ameyaw et al. 2024<sup>75</sup> | Option B+ | 176 | 176 | 100 |
| Azanaw et al. 2023<sup>77</sup> | Option B+ | 403 | 403 | 100 |
| Camara et al. 2024<sup>79</sup> | Option B+ | 5467 | 5467 | 100 |
| Amone et al. 2024<sup>89</sup> | Option B+ | 270 | 270 | 100 |
| Greenberg et al. 2025<sup>81</sup> | Option B+ | 304 | 304 | 100 |
| Mutale et al. 2025<sup>82</sup> | Option B+ | 49 | 49 | 100 |
| Njah et al. 2023<sup>83</sup> | Option B+ | 560 | 560 | 100 |
| Odongo et al. 2023<sup>90</sup> | Option B+ | 368 | 368 | 100 |
| Bengtson et al. 2024<sup>91</sup> | Option B+ | 399 | 402 | 99 |
| Young et al. 2024<sup>86</sup> | Option B+ | 1497 | 1535 | 98 |
| Phelanyane et al. 2023<sup>92</sup> | Option B+ | 2251 | 2549 | 88 |
| Tenthani et al. 2014<sup>58</sup> | Option B+ | 10179 | 11534 | 88 |
| Tolossa et al. 2020<sup>87</sup> | Option B+ | 287 | 330 | 87 |
| Phiri et al. 2017<sup>63</sup> | Option B+ | 1082 | 1269 | 85 |
| Reidy et al. 2019<sup>60</sup> | Option B+ | 357 | 434 | 82 |
| Schwartz et al. 2015<sup>59</sup> | Option B+ | 77 | 100 | 77 |
| Etoori et al. 2018<sup>43</sup> | Option B+ | 496 | 665 | 75 |
| Phillips et al. 2024<sup>93</sup> | Option B+ | 4559 | 6680 | 68 |
| Atanga et al. 2017<sup>37</sup> | Option B+ | 268 | 404 | 66 |
| Kim et al. 2015<sup>19</sup> | Option B+ | 759 | 1417 | 54 |
| Chan et al. 2016<sup>39</sup> | Option B+ | 456 | 914 | 50 |
| Dzangare et al. 2016<sup>41</sup> | Option B+ | 138 | 282 | 49 |
| Slogrove et al. 2024<sup>78</sup> | Option B+ | 9276 | 32015 | 29 |

### Retention in HIV care

Our second review outcome was proportion of women retained in HIV care. In total, 31 studies (38%) reported proportion retained in care (Supplementary Table 1) among pregnant and postpartum women with HIV in sub-Saharan Africa. Six and 12 months were the two most commonly reported time periods. The mean estimate of retention in HIV care prior to Option B+ was 44% versus 64% after implementation of Option B+ (RR=1.46, 95% CI=1.41-1.51).

Figure 3 illustrates results of the data synthesis and meta-analysis. Overall, pooled estimates of retention in care were 66.5% (95% CI: 59.0%, 73.9%) at 6 months (Figure 2a) (I^2^>99.8%) and 64.1% (95% CI: 57.3%, 71.0%) at 12 months (Figure 2b), (I^2^>99.7%) showing high degree of heterogeneity at both time points. At 6 months post-ART initiation, the pooled estimates for retention in care were 36.9% (95% CI: 13.9%, 59.9%) before the implementation of Option B+ and 72.7% (95% CI: 66.3%, 79.1%) after implementation. At 12 months post-ART initiation, retention rates were 64.8% (95% CI: 57.5%, 72.0%) prior to Option B+ and 64.0% (95% CI: 51.8%, 65.8%) following its implementation.

**Figure 3:**
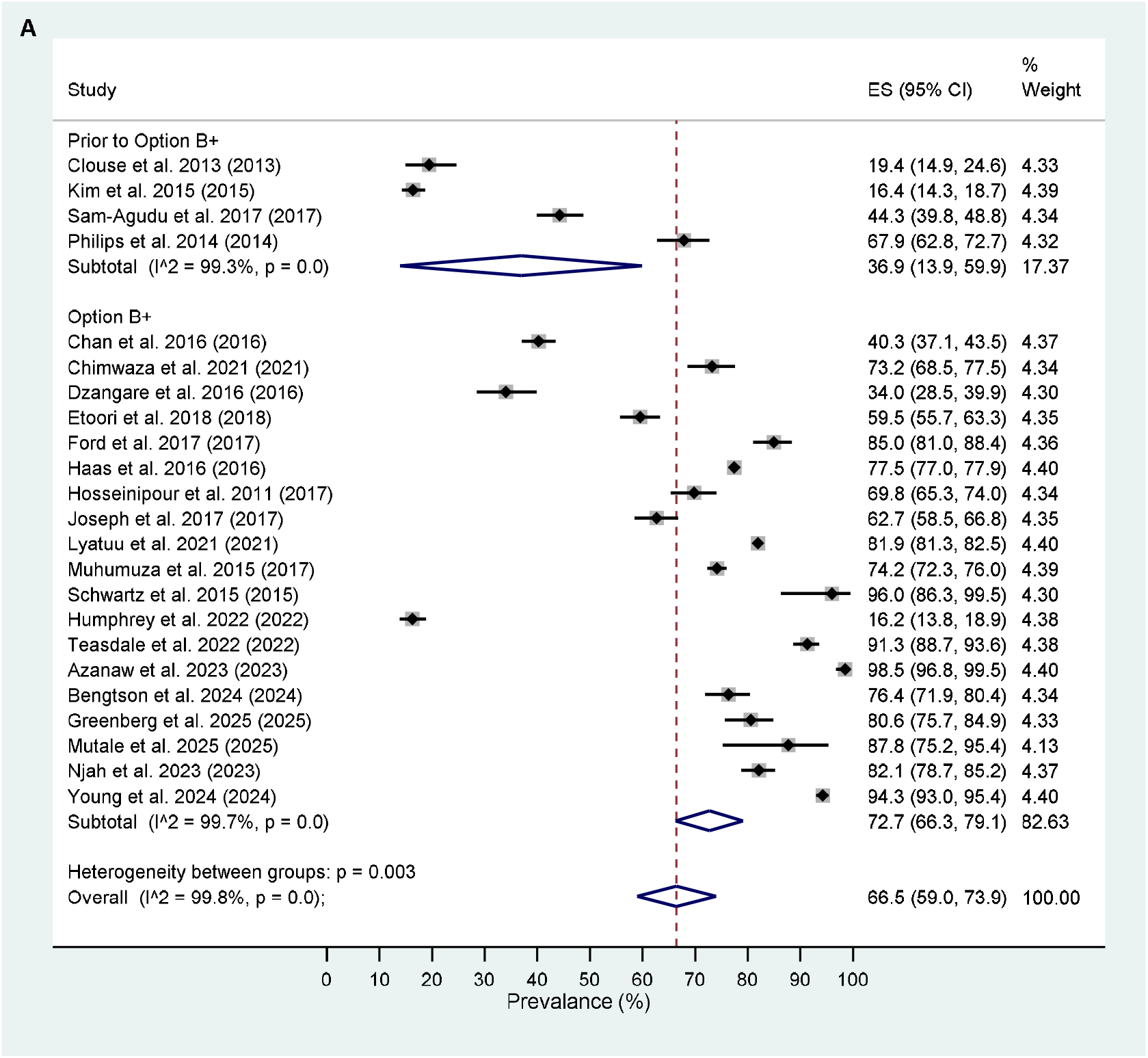
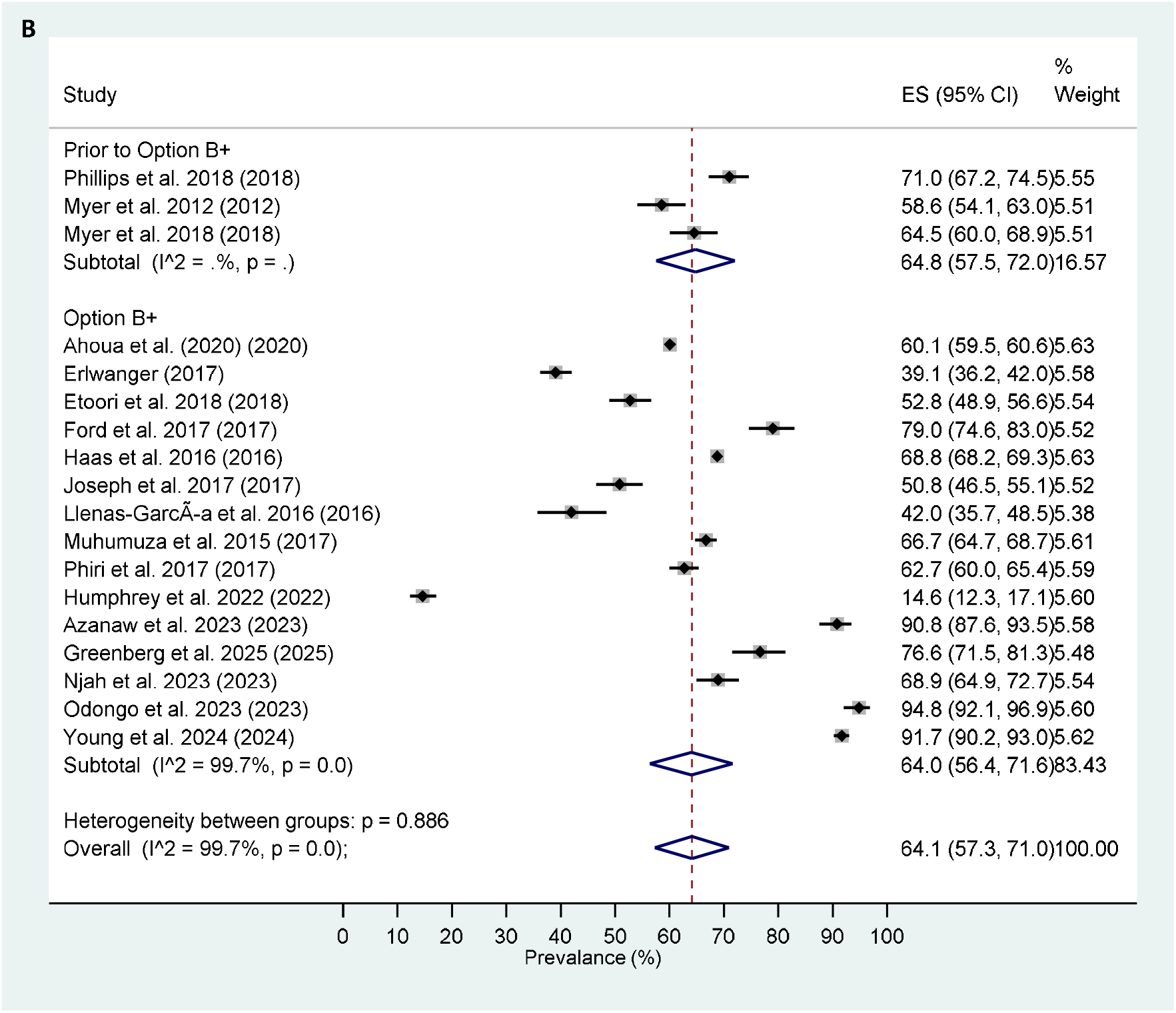
Pooled estimates of (A) 6-month and (B) 12-month retention among pregnant women.

### Critical appraisal and risk of bias within studies

The critical appraisal and risk of bias evaluation utilized the Joanna Briggs Institute (JBI) critical appraisal checklist designed for studies reporting prevalence data, using a scale where 1 is lowest and 9 is highest. Each appraisal point was systematically reviewed, and results are presented in Supplementary File 2. Median score across all studies was 7 out of 9 points (IQR: 6-8) with results ranging from 1 to 9.

## DISCUSSION

This systematic review sought to synthesize the literature on the uptake of HIV care and retention in HIV care among pregnant and postpartum women before and after Option B+ implementation, and to assess the impact of these policies on continuity of HIV care. As countries make progress towards ending the HIV epidemic, universal access to HIV care and sustained retention through successful and sustained viral suppression remain the key goals.^94^ These goals are of particular importance in pregnant women where achieving viral suppression is critical not only for maternal health and treatment outcomes but also to prevent vertical transmission of HIV to the child. Sub-Saharan African countries remain among those with the largest burden of HIV among women,^95^ including estimates of 13% in Zimbabwe, 21% in Botswana, 23% in South Africa, 24% in Lesotho, and 30% in Eswatini.^96^ Prevalence is particularly high among pregnant women in the region, with estimates of 28% in South Africa,^97^ 17% in Zimbabwe,^98^ and 22% in Botswana.^99^ Pregnancy and the postpartum period are known to be periods of high risk for disengagement from HIV care and treatment for many reasons, including mobility after delivery.^64, 23,100^

Our review of 82 articles identified some important findings. First, implementation of Option B+ was associated with higher rates of initiation of ART among pregnant women. The pooled analysis indicated an increase in the median uptake of HIV care from 90% before the implementation of Option B+ to 100% after implementation. Although studies may have been biased by including only women who initiated ART, more than 60% of the studies included reported a 100% uptake of HIV care, highlighting the impact of the Option B+ strategy. Furthermore, the analysis estimated an 8-percentage point increase in likelihood of initiating ART compared to the period prior to Option B+ implementation. These findings support the conclusion that Option B+ is associated with enhanced access to HIV care among pregnant women and emphasizes the importance of continued support and implementation of such strategies to further improve maternal HIV care outcomes. Though our results do not directly assess the reasons for the improvement in uptake of HIV services, the authors of the studies reviewed suggest proactive and trained healthcare providers, awareness of Option B+ in the community,^35^ male involvement, integration of mother and child services^25^ and raised awareness and educated people across the entire country about the programme^43^ as facilitators to programme success.

Second, retention in HIV care was higher after Option B+ implementation. The observed increase in median retention from 67% before Option B+ to 74% at 6 months after implementation highlights that the positive impact of the Option B+ strategy in the early postpartum period. Pooled estimates of retention in care were 67% at 6 months and 60% at 12 months in the articles reviewed. Our pooled retention estimates were lower than that reported in a 2018 review on HIV care initiation and retention for pregnant and postpartum women after starting ART after Option B+ implementation, Knettel et al. reported retention estimates of 80% at 6 months and 75% at 12 months.^101^ This difference in retention rates could be attributed to differences in the definition of retention between our study and Knettel et al. The decrease in median retention at 12 and 24 months suggests potential challenges in sustaining long-term engagement in care, warranting further investigation into factors influencing retention during the extended phases of HIV care. Our analysis also highlights that few studies report long-term outcomes, such as 24-months after delivery, and emphasize the need for longer-term retention explorations. Overall, these findings emphasize the effectiveness of the Option B+ strategy in enhancing short-term retention and call for ongoing efforts to address challenges associated with long-term retention in HIV care programs.

However, retention in HIV care among pregnant women remains below targets required to end the HIV epidemic. The median retention rate observed at 6 months was 74% with slightly lower estimates at 12 months and 24 months (69% and 59%, respectively). These findings among pregnant women were lower than among non-pregnant adults who initiated ART at public sector clinics in South Africa after January 2018, who had a retention of 77%, 12 months after ART initiation.^102^

Our results should be interpreted in light of some potential limitations in our approach. While our search terms were developed by an experienced research librarian, the terminology used to describe uptake of HIV services and retention on ART vary widely across the published literature and where our search strategy did not include specific terminology, we may have missed relevant articles. We searched grey literature and references of prior reviews to mitigate this risk. We conducted an Egger’s test and created a funnel plot to assess publication bias. The results indicated an over-representation of studies with significant or favorable findings. Consequently, the pooled estimates may overstate the effect of Option B+ implementation on both the uptake of HIV services and retention outcomes. This review includes only published studies, which may limit generalizability. Additionally, the included reviews had varying implementation timeframes and differing definitions of uptake and retention. Another limitation is the challenge of determining whether the observed results were directly caused by policy implementation or simply reflected existing trends. Finally, the review consists of studies that may have included only individuals who initiated ART, potentially leading to an overestimation of HIV care uptake proportions. Another limitation is that we only included English articles, thereby excluding some countries in West Africa. Despite this, our review included over 60 articles representing 15 countries in the sub-Saharan region over more than a decade. We reviewed articles before and after Option B+ implementation and in doing so, aimed to capture the most comprehensive estimates of outcomes across these time periods.

## CONCLUSION

Uptake and retention in HIV care for pregnant and postpartum women increased after implementation of Option B+. This emphasizes the importance of continued support and implementation of such strategies to further improve maternal HIV care outcomes. Retention rates also increased after Option B+ implementation but remain sub-optimal to achieve the 95-95-95 goals set by UNAIDS. Targeted interventions are needed to improve retention among pregnant and postpartum women living with HIV.

## Supporting information

Supplementary File 1. PROSPERO registration

Supplementary File 2. JBI Critical Appraisal Checklist.

## Data Availability

All data generated or analysed during this study are included in this published article.

## Author contributions

MM, CH and NJ conceptualized the study and methodology. CH, NJ, MB, NN, LR and CN performed the screening and extraction of data. NJ wrote the original draft. MM and NJ were responsible for statistical analysis. MM, LR and KC provided interpretation of results. All authors critically reviewed the manuscript.

## Conflict of interest statement

MDH has received travel support from the World Heart Federation and consulting fees from PwC Switzerland. MDH has an appointment at The George Institute for Global Health, which has a patent, license, and has received investment funding with intent to commercialize fixed-dose combination therapy through its social enterprise business, George Medicines. MDH has pending patents for heart failure polypills. The other authors declare that the research was conducted in the absence of any commercial or financial relationships that could be construed as a potential conflict of interest.

## Funding source

Research reported in this publication was supported by the National Institutes of Health (NIH) through the Eunice Kennedy Shriver National Institute of Child Health & Human Development and the National Institute for Allergy and Infectious Diseases under grant R01 HD103466, and the Fogarty International Center and National Institute of Mental Health under award number D43 TW010543. The content is solely the responsibility of the authors and does not necessarily represent the official views of the National Institutes of Health.

## Availability of data and materials

All data generated or analysed during this study are included in this published article.

## Supplementary Table

**Supplementary Table 1.**
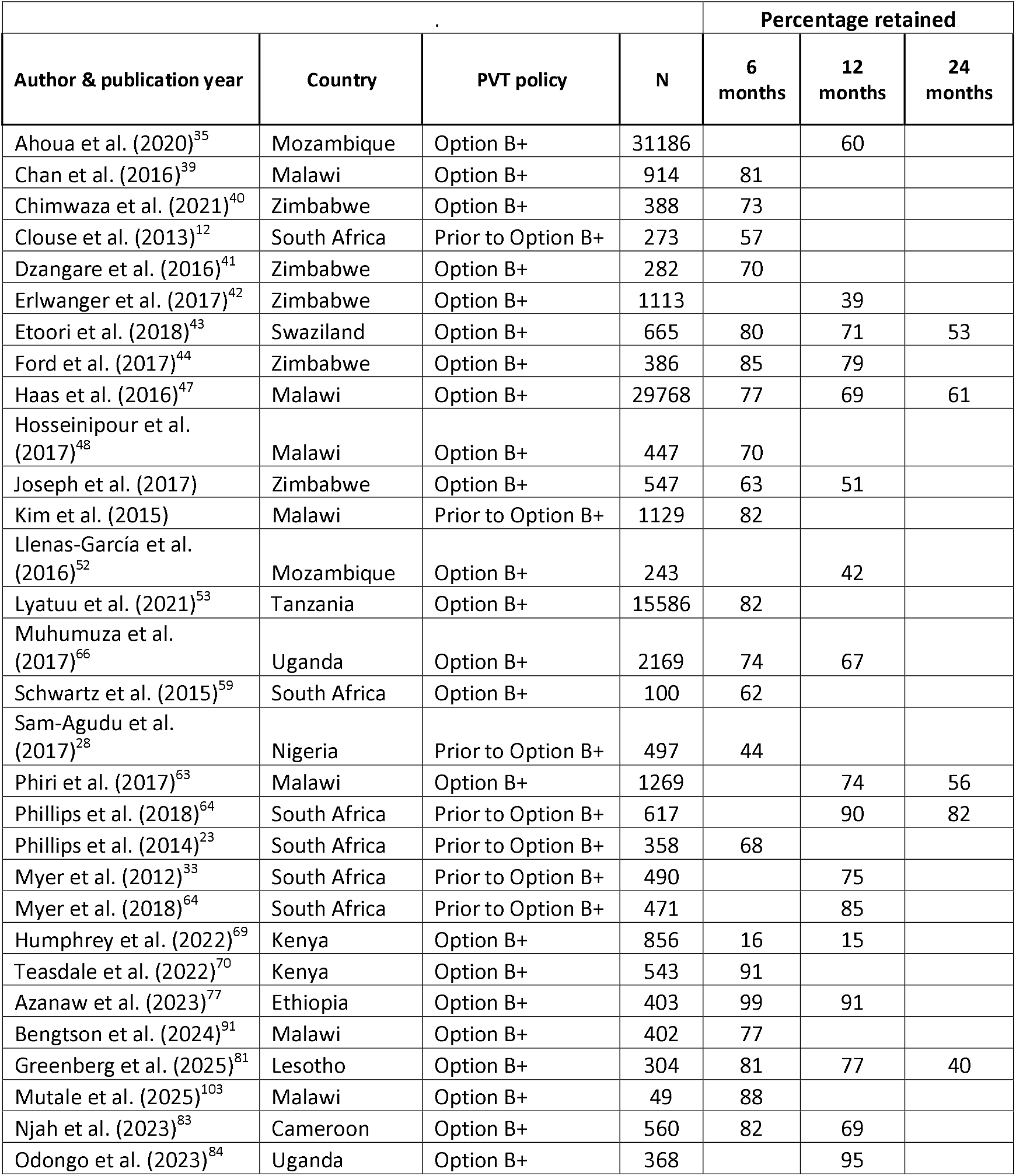
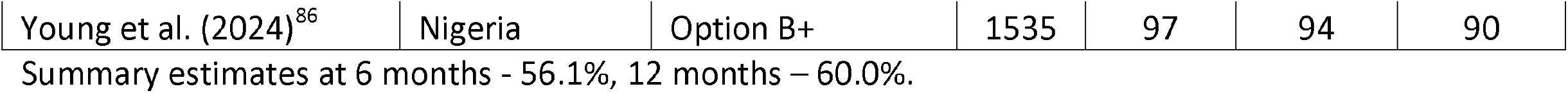
Retention in HIV care among pregnant women at 6-, 12- and 24 months follow up in a systematic review of studies of prevention of vertical transmission of HIV in sub-Saharan Africa from 2010-2023.

## Appendix

**Table 1.**
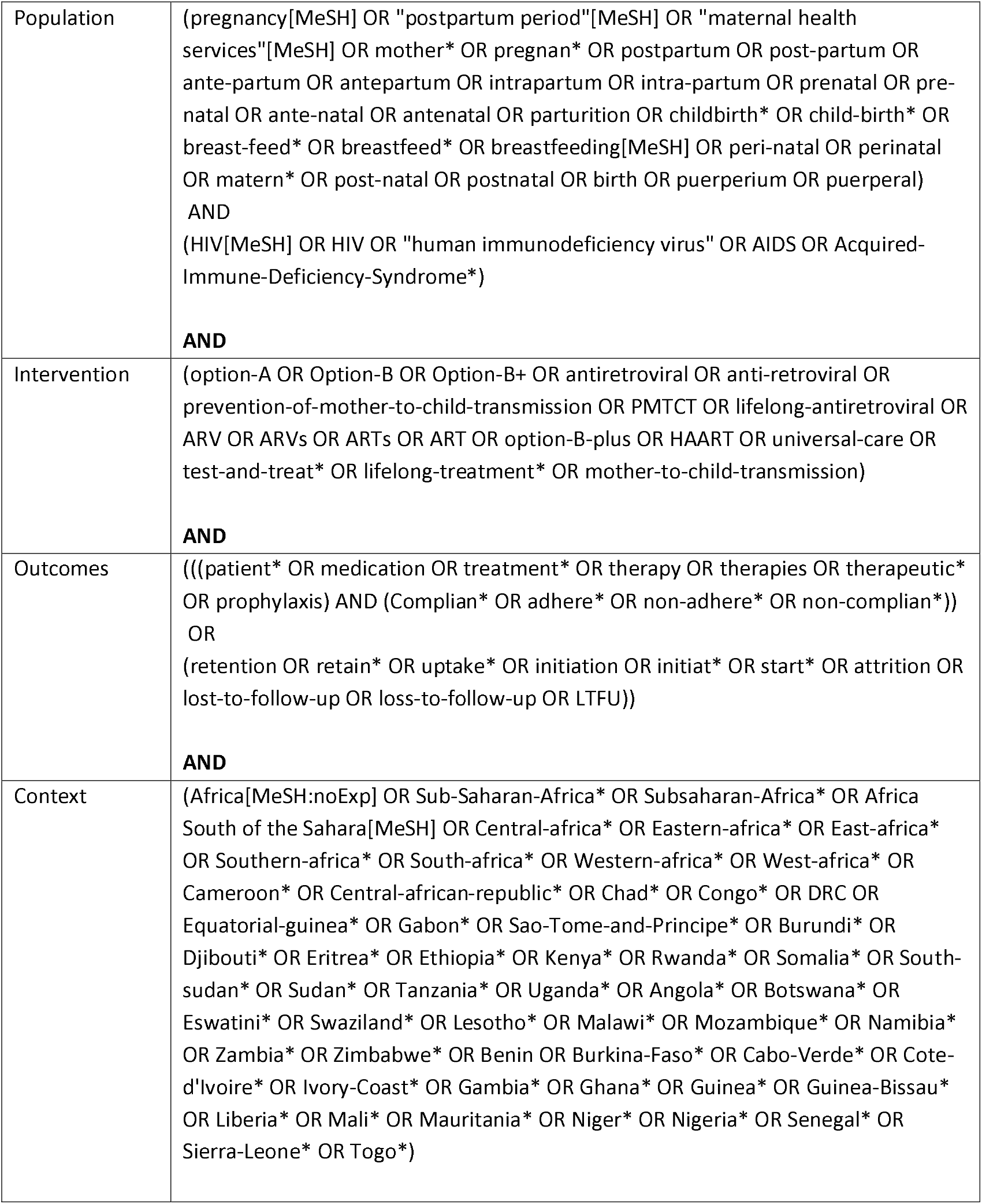
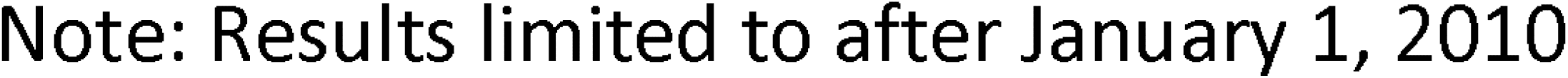
Search Strategy – PubMed.

## Notes

### Competing Interest Statement

The authors have declared no competing interest.

