## Supplementary File 1. PROSPERO registration for "Uptake and retention in HIV care among pregnant and postpartum women living with HIV under different eras of vertical transmission prevention policies in sub-Saharan Africa: a systematic review and meta-analysis"

PROSPERO
International prospective register of systematic reviews

### Uptake and retention in HIV care among pregnant and postpartum women living with HIV under different eras of prevention of mother-to-child transmission (PMTCT) policies in sub-Saharan Africa: a systematic review and meta-analysis

*Candice Hwang, Cornelius Nattey, Bernard Mbwele, Nkosinathi Ngcobo, Molly Beestrum, Mark D. Huffman, Matthew Fox, Kate Clouse, Mhairi Maskew*

To enable PROSPERO to focus on COVID-19 submissions, this registration record has undergone basic automated checks for eligibility and is published exactly as submitted. PROSPERO has never provided peer review, and usual checking by the PROSPERO team does not endorse content. Therefore, automatically published records should be treated as any other PROSPERO registration. Further detail is provided **here**.

##### Citation

Candice Hwang, Cornelius Nattey, Bernard Mbwele, Nkosinathi Ngcobo, Molly Beestrum, Mark D. Huffman, Matthew Fox, Kate Clouse, Mhairi Maskew. Uptake and retention in HIV care among pregnant and postpartum women living with HIV under different eras of prevention of mother-to-child transmission (PMTCT) policies in sub-Saharan Africa: a systematic review and meta-analysis. PROSPERO 2021 CRD42021290122 Available from: https://www.crd.york.ac.uk/prospero/display_record.php?ID=CRD42021290122

##### Review question

What are the outcomes related to the (a) uptake and (b) retention in HIV care among pregnant and postpartum women living with HIV with implementation of different PMTCT policies (Option A, B, or B+) in sub-Saharan Africa?

##### Searches

We will search the following databases: PubMed; Embase; Cochrane Library; Scopus; African Index Medicus; and abstracts from the International AIDS Conference, International AIDS Society (IAS) Conference on HIV Science, and Conference on Retroviruses and Opportunistic Infections (CROI). Searches will be limited to English language publications. Results will be limited to those published after Jan 1, 2010, because the WHO recommendations that introduced Option A and B were published in 2010. Results up to the date of search will be included. If over twelve months pass between the initial search and data analysis, then an additional search will be performed to cover the intervening time.

##### Types of study to be included

Included studies can be cohort, case control, cross-sectional, or interventional studies. In interventional studies where retention or uptake is the target of intervention, only data from the control arm will be included. White papers, commentaries, modeling studies, cost-effectiveness studies, and case studies will be excluded. Studies can involve any type of facility or provider. Qualitative studies will be excluded unless there is specific data on update or retention outcomes. Existing systematic reviews will be evaluated for additional, non-duplicate references. Published protocols with no data will be excluded, but if the study meets the inclusion criteria, then we will attempt to find study results to evaluate for inclusion.

**Condition or domain being studied**

Uptake or retention in HIV care outcomes for pregnant and post-partum women in the context of a PMTCT policy implemented in the country.

**Participants/population**

Pregnant and postpartum women living with HIV whose uptake or retention in HIV care has been measured in any sub-Saharan African country.

**Intervention(s), exposure(s)**

Implementation of a PMTCT policy in the country, such as Option A, B, or B+.

**Comparator(s)/control**

A comparator is not applicable because included studies only need to measure the rate of uptake or retention in care. Studies are not required to complete an intervention, and therefore do not need to include a comparison group.

**Main outcome(s)**

Primary outcomes for uptake of care:
- Proportion of pregnant women on ART prior to initial ANC visit
- Proportion of pregnant women initiating ART during pregnancy

Primary outcome for retention in care:
- Proportion of women retained in HIV care, as defined by the study, after the initial ANC visit or after ART initiation, recorded at all available time points reported, with retention at 12 months as the primary outcome. Patients reported as non-adherent to ART but attending clinic appointments (or meeting other study-defined criteria for retention) will be counted as retained. Patients who transferred care are considered retained.

**Additional outcome(s)**

Secondary outcomes for uptake of care:
- Median gestational age at initiation of ART during pregnancy
- Median gestational age at initial ANC visit
- Mean CD4 count at initial ANC visit
- Mean viral load at initial ANC visit
- Mean viral load (or categorical suppressed vs not suppressed) at delivery

Secondary outcomes for retention in care include, at each available time point, proportion of women:
- Retained at original site
- Non-adherent to ART while continuing to attend clinic visits
- Formally transferred care
- Lost to follow up (discontinued visits, informal self-transfers, undocumented deaths)
- Known to have died

**Data extraction (selection and coding)**

References will be managed in EndNote, and duplicates will be removed. Rayyan will be used to support screening of studies and track inclusion or exclusion of studies. Two independent reviewers will be involved in each phase of review, including title and abstract screening, full-text screening, and data extraction. Title and abstract screening will include a pilot test to assess if the reviewers are able to achieve over 90% agreement on inclusion based on the eligibility criteria listed in this protocol. For all phases of review, CH will be one reviewer for each study, and the second reviewer will BM, CN, or NN, with studies randomly distributed among them. Disagreements will be resolved through discussion to achieve consensus or input from MM will be sought to achieve consensus. After full text screening, relevant non-duplicate citations from previous systematic reviews and other studies labelled for snowball in the initial search will be added for inclusion/exclusion consideration in a snowball phase. Data extraction will use a pre-determined template that captures all relevant variables. Data extraction will be done independently by two reviewers, with CH as one reviewer and BM, CN, or NN as the second reviewer, and discrepancies will be resolved by consensus.

Apart from outcomes of interest, data items to be extracted from studies include:
- Country
- District or locality
- Facility type or setting
- Study year(s)
- Length of patient follow-up
- PMTCT protocol(s) that were used
- Study sample size
- Patient characteristics, including: Mean age; Gravida; Risk factors for HIV infection, such as injection drug users and female sex workers; Clinical indicators of disease stage such as WHO stage; Other co-morbidities
- Study design (e.g., cohort, case control, cross-sectional, interventional)
- Descriptions of routine care, including whether the study was designed to influence uptake or retention

**Risk of bias (quality) assessment**

Risk of bias and quality of individual studies that are included in this systematic review will be evaluated with the Joanna Briggs Institute critical appraisal checklist most relevant to the study type, as most included studies will likely be observational. Risk of bias will be considered in overall confidence of synthesized outcomes.

**Strategy for data synthesis**

Uptake outcomes will be recorded as reported by individual studies. Retention outcomes will be recorded at 6, 12, 18, 24, 48, and 60 months, if reported by individual studies. If specific timepoints are not reported, then outcomes will be assigned to the closest timepoint to median follow-up for the specific study. Missing timepoints will be interpolated, assuming a linear decline between two timepoints.

Primary uptake and retention at 12 months outcomes from all included studies will be synthesized using meta-analysis, and will be stratified by Option A/B vs Option B+ era. This stratification is according to which PMTCT policy was in place at the time the study was conducted in that country, and not according to global calendar time. For example, a study conducted in 2015 in South Africa would be stratified to the Option B+ era, while a different study in Nigeria also conducted in 2015 would be stratified to the Option A/B era. Retention at 6 and 24 months will also be summarized using meta-analysis without policy era stratification. Uptake and retention estimates and their 95% confidence intervals will be plotted using forest plots. Heterogeneity will be assessed with the $I^2$ statistic and is expected to be moderate to high. The pooled estimates will account for this heterogeneity using a random effects regression with a Freeman and Tukey arcsine transformation.

The expected heterogeneity will be investigated through random effects meta-regression of study-level variables, including study year, mean CD4 count at initial ANC visit, median gestational age at initial ANC visit, and either mean maternal age or gravida depending on which is more widely reported. Meta-regression will be presented as a scatterplot of the relationship of the primary uptake and retention at 12-months outcomes against the study-level variable of interest. The degree of association and statistical significance will be reported.

**Analysis of subgroups or subsets**

Uptake and retention at 12 months will also be summarized by geography, through weighted averages of the outcomes. If published studies are clustered in specific countries, then the summary data will be presented by country. Otherwise, it will be presented by region of Africa: Eastern, Central, Southern, and Western Africa.

We will also explore limitations in estimating uptake and retention outcomes, including the effect of including data from the control arm of interventional studies and whether there is bias in individual studies' length of follow-up based on retention outcomes. Weighted averages of primary uptake and retention outcomes from the control arm of interventional studies will be compared to that from observational studies because the increased resources associated with interventional studies may improve outcomes in the control arm over routine care in observational studies. Furthermore, for retention, individual studies could choose to stop reporting after a shorter amount of follow-up time if their cohorts have worse retention. We will explore this by comparing average retention at the last common time point used for reporting among studies with shorter vs longer follow-up.

Finally, summary data on primary and secondary outcomes for uptake and retention will be compared to UNAIDS 2025 targets. Specifically, we are interested in the targets of having 90% of women living with HIV on ART before their current pregnancy, 95% of pregnant women living with HIV achieve viral suppression before delivery, and 95% of breastfeeding women living with HIV achieve viral suppression at 6-12 months postpartum.

**Contact details for further information**

Candice Hwang


**Organisational affiliation of the review**

Health Economics and Epidemiology Research Office, Department of Internal Medicine, School of Clinical Medicine, Faculty of Health Sciences, University of the Witwatersrand, Johannesburg, South Africa.
https://www.heroza.org/

**Review team members and their organisational affiliations**

Ms Candice Hwang. Health Economics and Epidemiology Research Office, Department of Internal Medicine, School of Clinical Medicine, Faculty of Health Sciences, University of the Witwatersrand, Johannesburg, South Africa; Northwestern University Feinberg School of Medicine, Chicago, Illinois, USA.
Mr Cornelius Nattey. Health Economics and Epidemiology Research Office, Department of Internal Medicine, School of Clinical Medicine, Faculty of Health Sciences, University of the Witwatersrand, Johannesburg, South Africa.
Dr Bernard Mbwele. Department of Epidemiology, University of Dar es Salaam Mbeya College of Health and Allied Sciences, Mbeya, Tanzania.
Mr Nkosinathi Ngcobo. Health Economics and Epidemiology Research Office, Department of Internal Medicine, School of Clinical Medicine, Faculty of Health Sciences, University of the Witwatersrand, Johannesburg, South Africa.
Ms Molly Beestrum. Galter Health Sciences Library, Northwestern University Feinberg School of Medicine, Chicago, Illinois, USA.
Dr Mark D. Huffman. Department of Preventive Medicine, Northwestern University Feinberg School of Medicine, Chicago, Illinois, USA; Division of Cardiology, Department of Medicine, Northwestern University Feinberg School of Medicine, Chicago, Illinois, USA; The George Institute for Global Health, University of New South Wales, Sydney, Australia.

Dr Matthew Fox. Health Economics and Epidemiology Research Office, Department of Internal Medicine, School of Clinical Medicine, Faculty of Health Sciences, University of the Witwatersrand, Johannesburg, South Africa; Department of Global Health, Boston University School of Public Health, Boston, Massachusetts, USA; Department of Epidemiology, Boston University School of Public Health, Boston, Massachusetts, USA.
Dr Kate Clouse. Health Economics and Epidemiology Research Office, Department of Internal Medicine, School of Clinical Medicine, Faculty of Health Sciences, University of the Witwatersrand, Johannesburg, South Africa; Vanderbilt University School of Nursing, Nashville, Tennessee, USA; Vanderbilt Institute for Global Health, Vanderbilt University Medical Center, Nashville, Tennessee, USA.
Dr Mhairi Maskew. Health Economics and Epidemiology Research Office, Department of Internal Medicine, School of Clinical Medicine, Faculty of Health Sciences, University of the Witwatersrand, Johannesburg, South Africa.

**Type and method of review**

Meta-analysis, Systematic review

**Anticipated or actual start date**

11 October 2021

**Anticipated completion date**

25 February 2022

**Funding sources/sponsors**

Research reported in this publication was supported by the National Institutes of Health (NIH) through the Fogarty International Center and National Institute of Mental Health under Award Number D43 TW010543; and by the Eunice Kennedy Shriver National Institute of Child Health & Human Development and the National Institute for Allergy and Infectious Diseases under grant R01 HD103466. The content is solely the responsibility of the authors and does not necessarily represent the official views of the National Institutes of Health.

***Grant number(s)***

***State the funder, grant or award number and the date of award***

NIH Fogarty International Center and National Institute of Mental Health: D43 TW010543
NIH Eunice Kennedy Shriver National Institute of Child Health & Human Development and the National Institute for Allergy and Infectious Diseases: R01 HD103466

**Conflicts of interest**

**Language**

English

**Country**

South Africa, Tanzania, United States of America

**Published protocol**

https://www.crd.york.ac.uk/PROSPEROFILES/290122_PROTOCOL_20211108.pdf

**Stage of review**

Review Ongoing

##### Subject index terms status

Subject indexing assigned by CRD

##### Subject index terms

Africa South of the Sahara; Female; HIV Infections; Humans; Infectious Disease Transmission, Vertical; Policy; Postpartum Period; Pregnancy

##### Date of registration in PROSPERO

09 December 2021

##### Date of first submission

08 November 2021

##### Stage of review at time of this submission

| Stage | Started | Completed |
|---|---|---|
| Preliminary searches | Yes | Yes |
| Piloting of the study selection process | Yes | Yes |
| Formal screening of search results against eligibility criteria | No | No |
| Data extraction | No | No |
| Risk of bias (quality) assessment | No | No |
| Data analysis | No | No |

*The record owner confirms that the information they have supplied for this submission is accurate and complete and they understand that deliberate provision of inaccurate information or omission of data may be construed as scientific misconduct.*

*The record owner confirms that they will update the status of the review when it is completed and will add publication details in due course.*

##### Versions

09 December 2021
09 December 2021

PROSPERO
This information has been provided by the named contact for this review. CRD has accepted this information in good faith and registered the review in PROSPERO. The registrant confirms that the information supplied for this submission is accurate and complete. CRD bears no responsibility or liability for the content of this registration record, any associated files or external websites.
