## Supplementary File 2. JBI Critical Appraisal Checklist. for "Uptake and retention in HIV care among pregnant and postpartum women living with HIV under different eras of vertical transmission prevention policies in sub-Saharan Africa: a systematic review and meta-analysis"

| Author/year | Checklist items | | | | | | | | |
|---|---|---|---|---|---|---|---|---|---|
| | 1. Was the sample frame appropriate to address the target population? | 2. Were study participants sampled in an appropriate way? | 3. Was the sample size adequate? | 4. Were the study subjects and the setting described in detail? | 5. Was the data analysis conducted with sufficient coverage of the identified sample? | 6. Were valid methods used for the identification of the condition? | 7. Was the condition measured in a standard, reliable way for all participants? | 8. Was there appropriate statistical analysis? | 9. Was the response rate adequate, and if ût, was the low response rate managed appropriately? |
| Yitayew et al. (2019) | ✓ | ✓ | ✓ | ✓ | ✓ | ✓ | ✓ | ✓ | ✗ |
| Abrams et al. (2019) | ✓ | ✓ | ✓ | ✗ | ✓ | ✓ | ✓ | ✓ | ✓ |
| Ahoua et al. (2020) | ✓ | ✓ | ✓ | ✗ | ✓ | ✓ | ✗ | ✓ | ✓ |
| Ahoua et al. (2020) | ✓ | ✓ | ✓ | ✓ | ✓ | ✓ | ✓ | ✓ | ✓ |
| Akama et al. (2019) | ✓ | ✓ | ✗ | ✗ | ✓ | ✓ | ✓ | ✓ | ✓ |
| Aliyu et al. (2016) | ✓ | ✓ | ✓ | ✓ | ✗ | ✓ | ✓ | ✓ | ✗ |
| Aliyu et al. (2015) | ✓ | ✓ | ✗ | ✓ | ✓ | ✓ | ✓ | ✓ | ✓ |
| Altan et al. (2016) | ✓ | ✗ | ✗ | ✗ | ✓ | ✓ | ✗ | ✗ | ✗ |
| Arreskov et al. (2010) | ✓ | ✗ | ✗ | ✗ | ✗ | ✗ | ✗ | ✗ | ✗ |
| Atanga et al. (2017) | ✓ | ✓ | ✗ | ✓ | ✓ | ✓ | ✓ | ✓ | ✓ |
| Bancheû et al. (2010) | ✗ | ✗ | ✓ | ✗ | ✓ | ✓ | ✗ | ✗ | ✗ |
| Bannink-Mbazzi et al. (2013) | ✓ | ✓ | ✓ | ✗ | ✓ | ✓ | ✓ | ✓ | ✓ |
| Chaka et al. (2019) | ✓ | ✗ | ✗ | ✗ | ✓ | ✓ | ✓ | ✓ | ✗ |
| Chan et al. (2016) | ✓ | ✓ | ✓ | ✗ | ✓ | ✓ | ✓ | ✓ | ✗ |
| Chimwaza et al. (2021) | ✓ | ✓ | ✓ | ✓ | ✓ | ✓ | ✓ | ✓ | ✗ |

| | | | | | | | | | |
|---|---|---|---|---|---|---|---|---|---|
| Clouse et al. (2013) | ✓ | ✗ | ✗ | ✓ | ✗ | ✓ | ✓ | ✓ | ✓ |
| Dinh et al. (2018) | ✓ | ✓ | ✓ | ✗ | ✓ | ✗ | ✓ | ✓ | ✓ |
| Dzangare et al. (2016) | ✓ | ✓ | ✓ | ✗ | ✓ | ✓ | ✗ | ✓ | ✓ |
| Erlwanger et al. (2017) | ✓ | ✓ | ✓ | ✓ | ✓ | ✓ | ✓ | ✓ | ✓ |
| Escamilla et al. (2015) | ✗ | ✗ | ✓ | ✓ | ✗ | ✓ | ✗ | ✓ | ✗ |
| Etoori et al. (2018) | ✓ | ✓ | ✓ | ✗ | ✓ | ✓ | ✓ | ✓ | ✓ |
| Fatti et al. (2016) | ✓ | ✓ | ✓ | ✗ | ✗ | ✓ | ✓ | ✓ | ✗ |
| Fatt et al. (2014) | ✓ | ✓ | ✓ | ✗ | ✓ | ✓ | ✓ | ✓ | ✗ |
| Ford et al. (2018) | ✓ | ✓ | ✗ | ✗ | ✓ | ✓ | ✓ | ✓ | ✗ |
| Ford et al. (2017) | ✓ | ✓ | ✓ | ✗ | ✓ | ✓ | ✓ | ✓ | ✗ |
| Gamell et al. (2017) | ✓ | ✓ | ✓ | ✗ | ✗ | ✗ | ✗ | ✓ | ✗ |
| Girma et al. (2017) | ✓ | ✓ | ✓ | ✓ | ✗ | ✓ | ✓ | ✓ | ✗ |
| Giuliaû et al. (2016) | ✓ | ✗ | ✓ | ✓ | ✗ | ✗ | ✗ | ✓ | ✗ |
| Granato et al. (2016) | ✓ | ✓ | ✓ | ✗ | ✓ | ✓ | ✓ | ✓ | ✗ |
| Haas et al. (2016) | ✓ | ✓ | ✓ | ✗ | ✓ | ✓ | ✗ | ✓ | ✗ |
| Herce et al. (2015) | ✓ | ✓ | ✓ | ✗ | ✓ | ✓ | ✓ | ✓ | ✗ |
| Holmes et al. (2018) | ✓ | ✓ | ✓ | ✗ | ✓ | ✓ | ✗ | ✓ | ✗ |
| Hosseinipour et al. (2017) | ✓ | ✓ | ✓ | ✗ | ✓ | ✓ | ✓ | ✓ | ✗ |
| Hussain et al. (2011) | ✓ | ✓ | ✓ | ✗ | ✓ | ✓ | ✗ | ✓ | ✗ |
| Joseph et al. (2017) | ✓ | ✓ | ✓ | ✗ | ✗ | ✓ | ✓ | ✓ | ✗ |

| | | | | | | | | | |
|---|---|---|---|---|---|---|---|---|---|
| Kamuyango et al. (2014) | ✓ | ✓ | ✗ | ✗ | ✓ | ✓ | ✗ | ✓ | ✗ |
| Kim et al. (2015) | ✗ | ✓ | ✓ | ✗ | ✗ | ✗ | ✗ | ✓ | ✗ |
| Kiwanuka et al. (2018) | ✓ | ✓ | ✓ | ✓ | ✓ | ✓ | ✓ | ✓ | ✗ |
| Koole et al. (2014) | ✓ | ✓ | ✓ | ✗ | ✓ | ✓ | ✗ | ✓ | ✗ |
| Landes et al. (2021) | ✓ | ✓ | ✓ | ✗ | ✓ | ✓ | ✓ | ✓ | ✓ |
| Llenas-GarcÃa et al. (2016) | ✓ | ✓ | ✓ | ✗ | ✓ | ✓ | ✓ | ✓ | ✗ |
| Lyatuu et al. (2021) | ✓ | ✓ | ✓ | ✗ | ✓ | ✓ | ✗ | ✓ | ✓ |
| Mandewo et al. (2020) | ✓ | ✓ | ✓ | ✗ | ✗ | ✓ | ✗ | ✓ | ✗ |
| Mazuguni et al. (2019) | ✓ | ✗ | ✓ | ✓ | ✓ | ✓ | ✓ | ✓ | ✗ |
| Mitiku et al. (2016) | ✓ | ✓ | ✓ | ✓ | ✓ | ✓ | ✓ | ✓ | ✗ |
| Mnyani et al. (2020) | ✓ | ✓ | ✓ | ✗ | ✓ | ✓ | ✗ | ✓ | ✗ |
| Mnyani et al. (2020) | ✓ | ✓ | ✓ | ✗ | ✓ | ✓ | ✗ | ✓ | ✗ |
| Muhumuza et al. (2017) | ✓ | ✓ | ✓ | ✗ | ✓ | ✓ | ✓ | ✓ | ✗ |
| Musarandega et al. (2017) | ✓ | ✓ | ✓ | ✗ | ✓ | ✓ | ✗ | ✓ | ✗ |
| Myburgh et al. (2017) | ✓ | ✗ | ✗ | ✓ | ✗ | ✓ | ✗ | ✓ | ✗ |
| Myer et al. (2015) | ✓ | ✓ | ✓ | ✗ | ✓ | ✓ | ✗ | ✓ | ✗ |
| Myer et al. (2018) | ✓ | ✗ | ✓ | ✓ | ✓ | ✓ | ✗ | ✓ | ✗ |
| Myer et al. (2012) | ✓ | ✗ | ✓ | ✓ | ✓ | ✓ | ✗ | ✓ | ✗ |
| Vogt et al. (2015) | ✓ | ✓ | ✓ | ✗ | ✓ | ✓ | ✓ | ✓ | ✓ |
| Van Schalkwyk et al. (2013) | ✓ | ✓ | ✗ | ✗ | ✓ | ✓ | ✓ | ✓ | ✗ |

| | | | | | | | | | |
|---|---|---|---|---|---|---|---|---|---|
| van Lettow et al. (2018) | ✓ | ✓ | ✓ | ✗ | ✓ | ✓ | ✓ | ✓ | ✓ |
| Tweya et al. (2014) | ✓ | ✓ | ✓ | ✗ | ✓ | ✓ | ✓ | ✓ | ✗ |
| Turan et al. (2015) | ✓ | ✓ | ✓ | ✓ | ✓ | ✓ | ✓ | ✓ | ✗ |
| Tenthani et al. (2014) | ✓ | ✓ | ✓ | ✗ | ✓ | ✓ | ✓ | ✓ | ✓ |
| Schwartz et al. (2015) | ✓ | ✓ | ✓ | ✗ | ✗ | ✓ | ✓ | ✓ | ✗ |
| Sam-Agudu et al. (2017) | ✓ | ✓ | ✓ | ✓ | ✓ | ✗ | ✗ | ✓ | ✓ |
| Ross et al. (2018) | ✓ | ✗ | ✓ | ✗ | ✓ | ✓ | ✓ | ✓ | ✗ |
| Reidy et al. (2019) | ✓ | ✗ | ✓ | ✗ | ✗ | ✗ | ✗ | ✓ | ✗ |
| Pricilla et al. (2018) | ✓ | ✓ | ✓ | ✗ | ✓ | ✓ | ✓ | ✓ | ✓ |
| Price et al. (2014) | ✓ | ✓ | ✓ | ✗ | ✓ | ✓ | ✓ | ✓ | ✗ |
| Phiri et al. (2017) | ✓ | ✓ | ✓ | ✗ | ✓ | ✓ | ✓ | ✓ | ✗ |
| Phillips et al. (2018) | ✓ | ✓ | ✓ | ✓ | ✓ | ✓ | ✓ | ✓ | ✓ |
| Phillips et al. (2014) | ✓ | ✗ | ✓ | ✓ | ✓ | ✓ | ✓ | ✓ | ✓ |
| Parker et al. (2015) | ✓ | ✓ | ✓ | ✓ | ✓ | ✓ | ✓ | ✓ | ✓ |
| Abuogi et al. (2022) | ✓ | ✓ | ✓ | ✓ | ✓ | ✓ | ✓ | ✓ | ✓ |
| Geremew et al. (2023) | ✓ | ✓ | ✓ | ✓ | ✓ | ✓ | ✓ | ✓ | ✓ |
| Humphrey et al. (2022) | ✓ | ✓ | ✓ | ✗ | ✓ | ✓ | ✓ | ✓ | ✓ |
| Teasdale et al. (2022) | ✓ | ✓ | ✓ | ✗ | ✗ | ✓ | ✓ | ✓ | ✗ |
| Humphrey et al. (2024) | ✓ | ✓ | ✓ | ✓ | ✓ | ✓ | ✓ | ✓ | ✓ |
| Naidoo et al. (2023) | ✓ | ✓ | ✓ | ✓ | ✓ | ✓ | ✓ | ✓ | ✓ |

| | | | | | | | | | |
|---|---|---|---|---|---|---|---|---|---|
| Phelanyane et al. (2023) | ✓ | ✓ | ✓ | ✓ | ✓ | ✓ | ✓ | ✓ | ✓ |
| Ameyaw et al. (2024) | ✓ | ✓ | ✓ | ✓ | ✓ | ✓ | ✓ | ✓ | ✓ |
| Giovenco et al. (2025) | ✓ | ✓ | ✓ | ✗ | ✓ | ✓ | ✓ | ✓ | ✓ |
| Azanaw et al. (2023) | ✓ | ✓ | ✓ | ✗ | ✓ | ✓ | ✓ | ✓ | ✗ |
| Slogrove et al. (2024) | ✓ | ✓ | ✓ | ✓ | ✓ | ✓ | ✓ | ✓ | ✗ |
| Camara et al. (2024) | ✓ | ✓ | ✓ | ✓ | ✓ | ✓ | ✓ | ✓ | ✗ |
| Amone et al. (2024) | ✓ | ✓ | ✓ | ✗ | ✓ | ✓ | ✓ | ✓ | ✓ |
| Bengtson et al. (2024) | ✓ | ✓ | ✓ | ✓ | ✓ | ✓ | ✗ | ✓ | ✓ |
| Greenberg et al. (2025) | ✓ | ✓ | ✓ | ✓ | ✓ | ✓ | ✓ | ✓ | ✗ |
| Hwang et al. (2024) | ✓ | ✓ | ✓ | ✓ | ✓ | ✓ | ✓ | ✓ | ✗ |
| Mutale et al. (2025) | ✓ | ✗ | ✓ | ✓ | ✓ | ✓ | ✓ | ✓ | ✗ |
| Njah et al. (2023) | ✓ | ✗ | ✓ | ✓ | ✓ | ✓ | ✓ | ✓ | ✗ |
| Odongo et al. (2023) | ✓ | ✓ | ✓ | ✓ | ✓ | ✓ | ✓ | ✓ | ✗ |
| Phillips et al. (2024) | ✓ | ✓ | ✓ | ✓ | ✓ | ✓ | ✓ | ✓ | ✗ |
| Young et al. (2024) | ✓ | ✗ | ✗ | ✗ | ✓ | ✓ | ✓ | ✓ | ✗ |
| Ongaki et al. (201) | ✓ | ✗ | ✓ | ✗ | ✗ | ✓ | ✗ | ✓ | ✗ |
| Wanyenza et al. (2018) | ✓ | ✓ | ✓ | ✓ | ✓ | ✓ | ✓ | ✓ | ✗ |
| Tolossa et al. (2020) | ✓ | ✓ | ✓ | ✓ | ✓ | ✓ | ✓ | ✓ | ✓ |
